# Using Hawkes Processes to model imported and local malaria cases in near-elimination settings

**DOI:** 10.1101/2020.07.17.20156174

**Authors:** H. Juliette T. Unwin, Isobel Routledge, Seth Flaxman, Marian-Andrei Rizoiu, Shengjie Lai, Justin Cohen, Daniel J. Weiss, Swapnil Mishra, Samir Bhatt

## Abstract

Developing new methods for modelling infectious diseases outbreaks is important for monitoring transmission and developing policy. In this paper we propose using semi-mechanistic Hawkes Processes for modelling malaria transmission in near-elimination settings. Hawkes Processes are mathematical methods that enable us to combine the benefits of both statistical and mechanistic models to recreate and forecast disease transmission beyond just malaria outbreak scenarios. These methods have been successfully used in social media and earthquake modelling, but are not yet widespread in epidemiology. By using domain-specific knowledge, we can both recreate transmission curves for malaria in China and Swaziland and disentangle the proportion of cases which are imported from those that are community based.

## 1 Introduction

Modelling infectious disease transmission is an important tool for monitoring outbreaks and developing public policy to limit the spread of the disease. One common source of data available during these types of outbreaks are line lists, or case counts, from surveillance systems. These define the time at which patients are infected, along with other epidemiological information such as the sex, age and symptoms of the patient, locations they were infected or live and if they’ve travelled recently. An ideal model would combine all the information available from the line lists with disease specific mechanisms developed by experts of the disease to recreate case counts over time and accurately predict future behaviour.

Traditionally, SIR (Susceptible - Infected - Recovered) type models, such as the seminal Kermack-McKendrick model [1], or individual based models (for example [2] and [3]) have been used to model disease outbreaks. These methods encode well known disease specific mechanisms and can produce very good fits to data. However, they can require large amounts of data to produce these accurate fits, are cumbersome and computationally demanding to simulate from, difficult to forecast with and may make strong assumptions such as the homogeneity of the population. Therefore, there is scope to develop new methods and software to simulate outbreak behaviour. An alternative method proposed by Routledge et al. [4, 5] estimates temporal and spatial reproduction numbers by studying information diffusion processes in the form of network models, which reconstruct information transmission using known or inferred times of infection in a Bayesian framework [6]. These methods provide an adaptable framework to integrate multiple data types at different scales and identify missing data or external infection sources, but require very good data sets to accurately be able to predict from the models [6, 7].

SIR models can be linked to a well known statistical point process called Hawkes Processes [8], which we propose is a better alternative to model infectious disease outbreaks if the data is of high enough fidelity. These processes are semi-mechanistic, so give us the ability to encode disease specific information such as serial interval and incubation period, but are easier and computationally cheaper to simulate from and fit to data. Hawkes Processes model the intensity of infectious diseases by separating out contributions from exogenous and endogenous processes. The relative contributions of these two terms is disease specific and may have different levels of importance depending on the disease. The majority of transmission of Ebola is direct contact by human, and Kelly et al. [9] has recreated the Democratic Republic of Congo epicurve, or cases counts over time, using a Hawkes Process model with just an endogenous term. However, there is a real need to correctly parameterise the exogenous terms for diseases such as malaria in near-elimination settings and cholera to be able to accurately reproduce and predict the spread of the disease.

In this paper we focus on applying Hawkes Processes to malaria in near-elimination settings, where current models may not be especially well suited [4, 10]. In 2016, the World Health Organisation identified 21 countries with the potential to eliminate malaria by 2020; seven of these countries (Algeria, China, El Salvador, Iran, Malaysia, Paraguay, and Timor-Leste) have eliminated malaria since that list was published [11]. Since then, The Lancet Commission has published research by Feachem et al. [12] suggesting that malaria eradication within a generation is ambitious, achievable and necessary, but there needs to be an immediate, firm, global commitment to achieving such eradication by 2050. This involves developing new methods for modelling near-elimination settings, which can accurately capture the behaviour and help governments and public health organisations implement the best interventions to bring their countries closer to elimination.

Malaria is a complex disease to model, especially in low transmission settings, where the entomological inoculation rate (number of infected bites a person receives) varies greatly in space due to focal transmission and is potentially unstable due to sensitivity to heterogeneity in vector populations [4, 13, 14]. There are also inaccuracies in parasite prevalence rate estimations below 1-5% because a large sample size is necessary to accurately predict the proportion of the population with malaria [15]. We hypothesise that Hawkes Process models will help provide new insight into malaria transmission in these settings.

In this paper, we first introduce the traditional Hawkes Process and define the basic fitting and simulation algorithms. We then use our knowledge of malaria in near-elimination settings to tailor our exogenous and endogenous terms to best fit the data. Finally, we apply our methods to recreate and forecast from data sets in China and Swaziland.

## 2 Background

A uni-variate Hawkes Process is a self-exciting point process with a conditional intensity, *λ*(*t*), defined as:

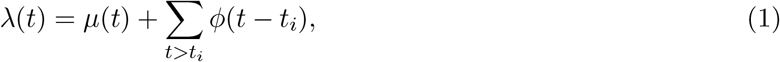

where *µ*(*t*) is the exogenous time dependent contribution to the intensity from external disease importations and 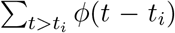 is the self exciting endogenous contribution representing person-to-person interactions [16]. Equation 1 means that the arrival of an event increases the likelihood of receiving a further event in the near future but that the importations are independent of all other events. Alternatively, a person getting infected increases the short term chance of other infections within the community, but people can also be infected independently from outside sources, such as zoonotic spillover or by travelling into the community already infected. The function *ϕ*(*·*) is often referred to as the triggering kernel in the Hawkes Process literature and describes a parameter similar to the serial interval distribution, or the expected time between infection and subsequent transmission. The parameter *t*_*i*_ refers to the times of the past events or in epidemiological applications, previous infections.

Similar to the simplest class of point processes, the Poisson Process [17], each event can be independently sampled from an intensity distribution. Unlike Poisson Processes, the intensity distribution of Hawkes processes is dependent on previous events because they are self-exciting, i.e. the occurrence of past events increases the likelihood of future events. The intensity of the Hawkes Processes is a stochastic function because it depends on event times which are random variables, however the Hawkes Process can be treated as a non-homogeneous Poisson Process between events. The methods have been used successfully to model earthquake [18], crime [19], financial time series [20] and social media [21, 22, 23, 24]. However, although a few people now use Hawkes Processes for epidemiological modelling [9, 25] they are not common place methods in this field yet.

The link between Susceptible - Infected - Recovered (SIR) and Hawkes Process models has been shown by Rizoiu et al. [8] for finite population sizes. They generalise the Hawkes Process to HawkesN and show that these types of models are conceptually similar to SIR models. The intensity function of HawkesN, *λ*_*N*_, is defined as

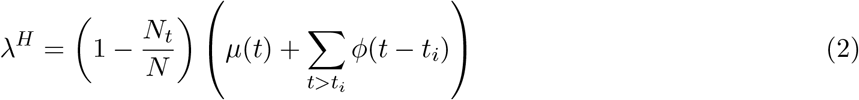

where *N* is the total population and *N*_*t*_ is the number of infected individuals. This is similar to the Hawkes Process intensity in equation (1) but also includes a population weighting term. Past events generate new events at a rate of *ϕ*(*t*) in HawkesN, which is analogous to the infection rate 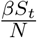 in the SIR models [1, 26], where *S*_*t*_ is the number of susceptible individuals at time *t*. Rizoiu et al. provide evidence that if the events in a HawkesN Process with parameters *{µ, α, δ, N}* have the intensity *λ*^*H*^(*t*) and the new infections of a stochastic SIR model with parameters *{β, γ, N}* (*γ* is the recovery rate) follow a point process of intensity *λ*^*I*^(*t*), the expectation of *λ*^*I*^(*t*) over all event times 𝒯 τ== = *τ*_1_, *τ*_2_, … is equal *λ*^*H*^(*t*):

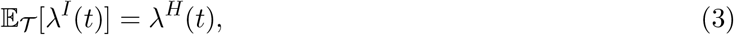

when *µ* = 0, *β* = *α* and *γ* = *θ*. In this paper we consider vanilla Hawkes Processes, instead of HawkesN, because we consider near-elimination malaria outbreaks where we assume an infinite susceptible population.

### 2.1 Fitting Hawkes Processes

We estimate the optimal parameters of our Hawkes Process, 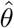, by minimising the negative log-likelihood function over our observed data

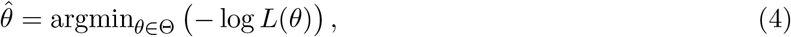

where

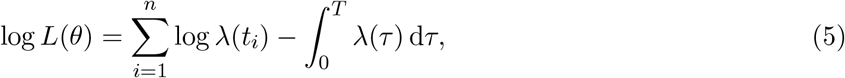

Θ is our parameter space and *n* is the number of events at time *T* [27]. For a full derivation see Appendix A.

Traditionally, a monotonically decreasing exponential kernel of the form

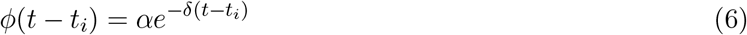

is used in the Hawkes process literature [9, 24, 28] where *δ* > *α* > 0. Here *α* controls the magnitude of the kernel, *δ* controls the speed of the decrease and *i* is an index. This kernel is traditionally chosen because in the most common use cases such as earthquakes and social media, events are most likely to trigger secondary events immediately after the first event happens. We discuss alternative kernels in Section 3 that may be better suited to epidemiological modelling, where for example latent periods are necessary to capturing disease specific behaviour.

### 2.2 Simulating Hawkes Processes

Simulation is used to learn more about our Hawkes Process so that we can better understand their behaviour and can validate our models to see how well they fit the underlying data. We can also use them to infer future behaviour. Ogata’s thinning algorithm [29] is a method for simulating non-homogeneous Poisson processes for any kernel function *ϕ*(*t*); we describe this algorithm adapted for Hawkes Processes in Algorithm 1.

#### Algorithm 1: Ogata’s thinning algorithm adapted for Hawkes Processes

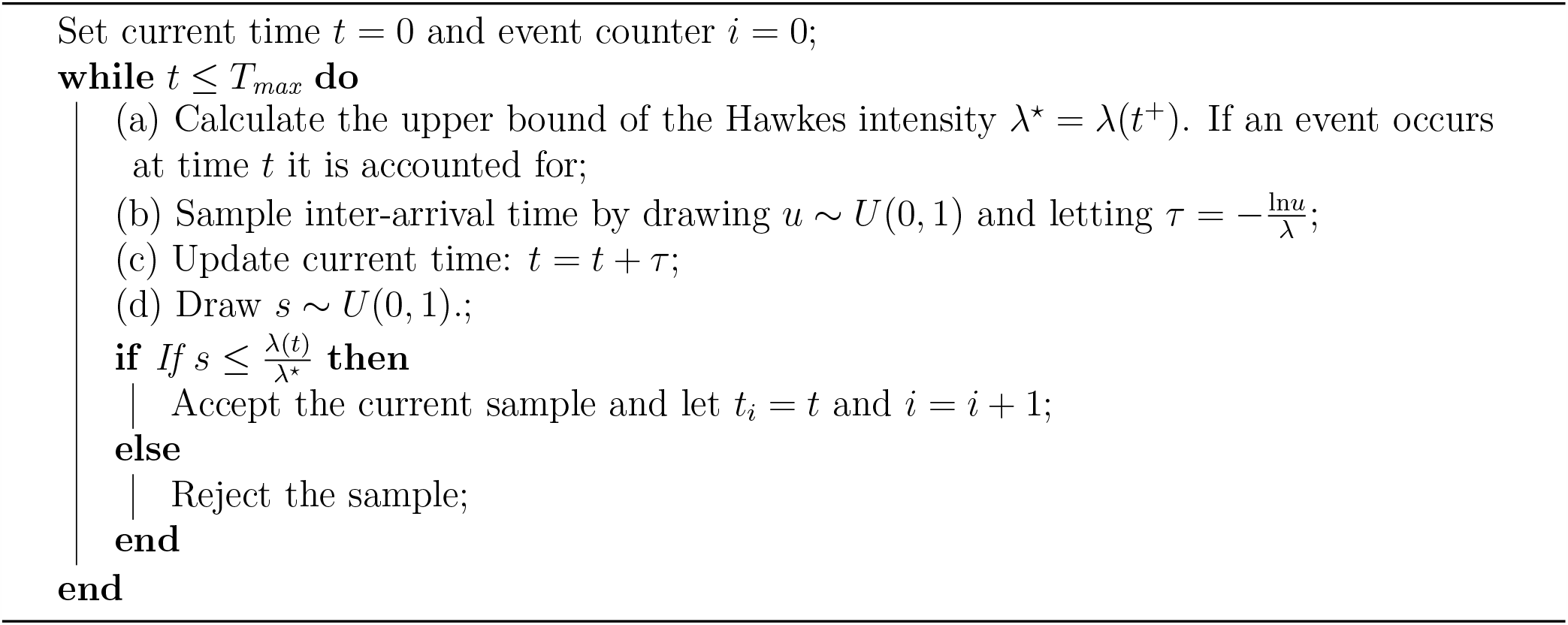

While the current time is less than the maximum time considered in the simulation, we calculate the maximum value of the intensity, *λ*^⋆^, for the events that have happened. For any bounded intensity *λ*(*t*), there is constant *λ*^⋆^ such that *λ*(*t*) ≤ *λ*^⋆^ in a given time interval. The upper bound of intensity is immediately after the event has occurred for a Hawkes Process with a monotonically decreasing kernel function, like the exponential function in equation 6, and no or a constant *µ* (exogenous term). However, this is not always so simple for other kernel functions and is addressed in Section 3.2.

Next we sample an inter-arrival time, *τ* ; the greater the maximum intensity, the higher the chance of subsequent infection arising and the shorter the suggested arrival time. This is then used to update the current time and the new event is then accepted or rejected according to *λ*^⋆^. If the event is accepted, the inter-arrival time is recorded and the event count incremented. Otherwise, we reject the inter-arrival time and repeat the sampling until one is accepted or the maximum time of the simulation is reached. Even if an inter-arrival time is rejected, the time counter is still updated [29]. The upper bound, *λ*^⋆^ is updated even in the case of a rejected inter-arrival time to improve efficiency because of the strict monotonicity of *λ*(*t*) in between event times.

## 3 Methods

Hawkes Processes are semi-mechanistic because we can incorporate disease specific information into our infection mechanism. Instead of using the traditional exponential kernel, we propose using a Rayleigh kernel of the form

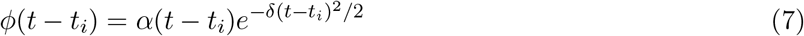

to model the within country transmission of malaria, where *α* > 0 controls the magnitude of the force of infection from an infected individual and *δ* > 0 controls the length of the infectious period. We choose this kernel because a person is not most infectious immediately after they are bitten by a mosquito. This kernel is little used in applications of Hawkes Process but has been suggested by Wallinga and Teunis [30], Gomez-Rodriguez et al. [31] and Ding et al. [32] and has already been used to represent the serial interval in malaria models [4]. We also used malaria domain specific knowledge to impose a delay between the person getting infected and becoming infectious, this is known as the latent period. Therefore, our kernel is

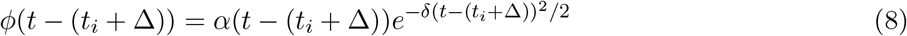

where Δ represents the delay and *t* > (*t*_*i*_ + Δ). We specify Δ = 12 days as done in literature [33]. This delay is novel and requires modifications to be made to Algorithm 1 because *λ*^⋆^ is no longer trivial to find; these are explained further in Section 3.2.

We also propose using a more complex time varying exogenous term than is found in literature (e.g. [23] and [28]) to capture the behaviour of the imported malaria cases. Our *µ* has the form

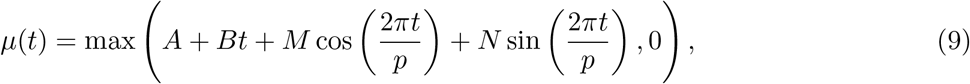

where *p* = 365.25 and *A, B, M* and *N* are constants that are fitted from data. This captures the linear decrease in exogenous events that we would expect in a malaria elimination setting along with the yearly fluctuating seasonality trends that often are associated with malaria. The *M* and *N* parameters will contribute less to the importations in areas with little or no seasonality. Unfortunately this also leads to a more complicated simulation process because the sinusoidal terms cause *µ* to increase periodically.

### 3.1 Fitting Hawkes Processes to malaria data

In this paper we fit our model to line lists of individuals with malaria in two countries over 1000 days. We consider *Plasmodium vivax* cases between 1^st^ January 2011 to 24^th^ September 2013 in Yunnan Province, China [5] and all malaria cases between 24^th^ January 2010 to 16^th^ November 2013 in Swaziland [34]. We chose these two data sets because the imported cases are labelled, although we do not use information about if a case was imported or local in our fitting process. Our cases are disaggregated by day, so we add right handed uniform jitter (ensuring the dates of each infection remain the same) to our times to ensure we have unique times for our events. This is a limitation of this method, but necessary for the Hawkes algorithm.

We use *optim* from the R stats package [35] to minimise our log-likelihood; the analytic directional derivatives of our log-likelihood are given in Appendix B. We initialise the optimisation routine from 100 different start points because our optimisation surface is complex and we often get stuck in local minima. This phenomenon is in part due to the comparatively small time period of observation for most disease time-series, when Hawkes Processes are defined until infinity, which means that the observed line lists could be recreated by a wide variety of parameter combinations. The likelihood loss function is also non convex [36]. We then select the two parameters which generate the minimum likelihood, perturb them 10 times and use them as starting points for the optimisation algorithm. Our final parameters are the ones with the minimum negative log-likelihood.

We can estimate the case reproduction number, *R*_*c*_ by considering the branching factor of the Hawkes Process. This is equal to the reproduction number in the presence of a range of interventions and is defined in Hawkes Process literature as the average number of children events that result from one parent event. This is derived in Appendix D for a Rayleigh kernel and is equal to the integral of the kernel between 0 and infinity:

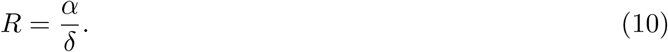

### 3.2 Simulating from a complex intensity function

It is no longer trivial to simulate from our intensity function for two reasons. First, our kernel is not monotonically decreasing and, second, we impose a fluctuating exogenous term. The maximum intensity from a single Rayleigh kernel at time *t* is

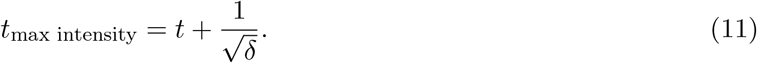

However, we can only place bounds on the time at which the intensity is maximum when it is comprised of multiple Rayleigh kernels, includes delays, *τ*, and has a time varying *µ*; we did not find an analytic solution. When *µ* = 0 or is constant, the maximum lies between *t*_last event_ and 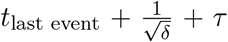, see Appendix C.1. These bounds have to be widened when considering non-monotonically decreasing exogenous terms because the maximum value of *λ* can occur after 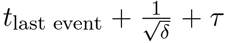 if *µ* periodically increases. In Figures 1A and B the maximum of the kernel still lies between the last event and the time of the maximum value of the kernel at that time. However, Figure C shows that the maximum value of lambda can occur outside that region and up until the maximum of the *µ* term. This is particularly important if the exogenous term dominates, which we predict happens in a near-elimination malaria settings.

**Figure 1:**
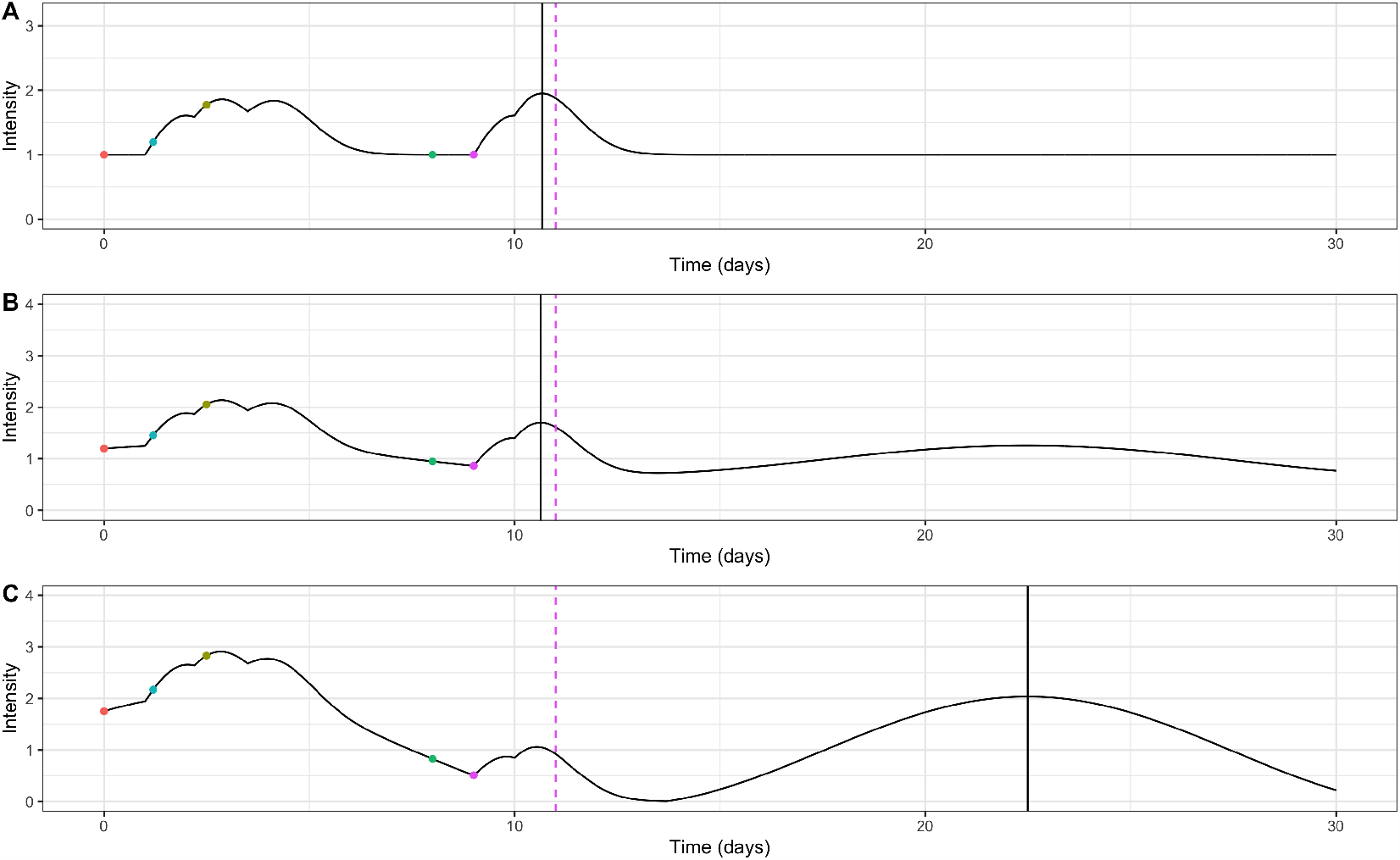
Illustrative plot of intensity function for events occurring at times 0, 1.2, 2.5, 8 and 9 with kernel parameters *α* = 1.0 and *δ* = 1.0, a 1 day delay and a time varying *µ*. The coloured dots refer to different events or infections and the dashed pink line indicate the time of the theoretical maximum value of a single Rayleigh kernel at the last event time. The solid black line indicates the time of the maximum value of the kernel after the last event. Figure A shows a constant *µ* and Figures B and C show sinusoidal *µ* with a linear decrease of different magnitudes. The parameters for equation (9) in each case are as follows: A - *A* = 1; B - *A* = 1, *B* = −0.001, *M* = 0.2, *N* = 0.2 and *p* = 20; C - *A* = 1, *B* = −0.001, *M* = 0.75, *N* = 0.75 and *p* = 20. These parameters are only illustrative and do not reflect parameters we would expect real in malaria models.

We propose a new algorithm for finding the maximum of *λ*(*t*). First we bound the times at which the maximum can occur; we calculate 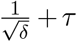 and the time of the maximum value of the exogenous term, *t*_*µ* max_ between the previous event and the final time of the simulation:

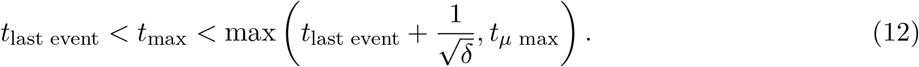

Since the intensity is the juxtaposition of multiple functions with known maximums, we can be sure that the maximum does not lie outside this bound. We then use a root finding algorithm similar to *uniroot*.*all* from the *rootSolve* package [37, 38] to locate all the roots of the derivative of the intensity. We do not know prior to the calculation how many roots there are so split the bound into a pre-defined number of sections and search for a sign change inside the interval. Once we have the times of these turning points, we evaluate them and find the maximum value of the intensity. This is summarised in Algorithm 2.

#### Algorithm 2: Algorithm for finding λ^⋆^

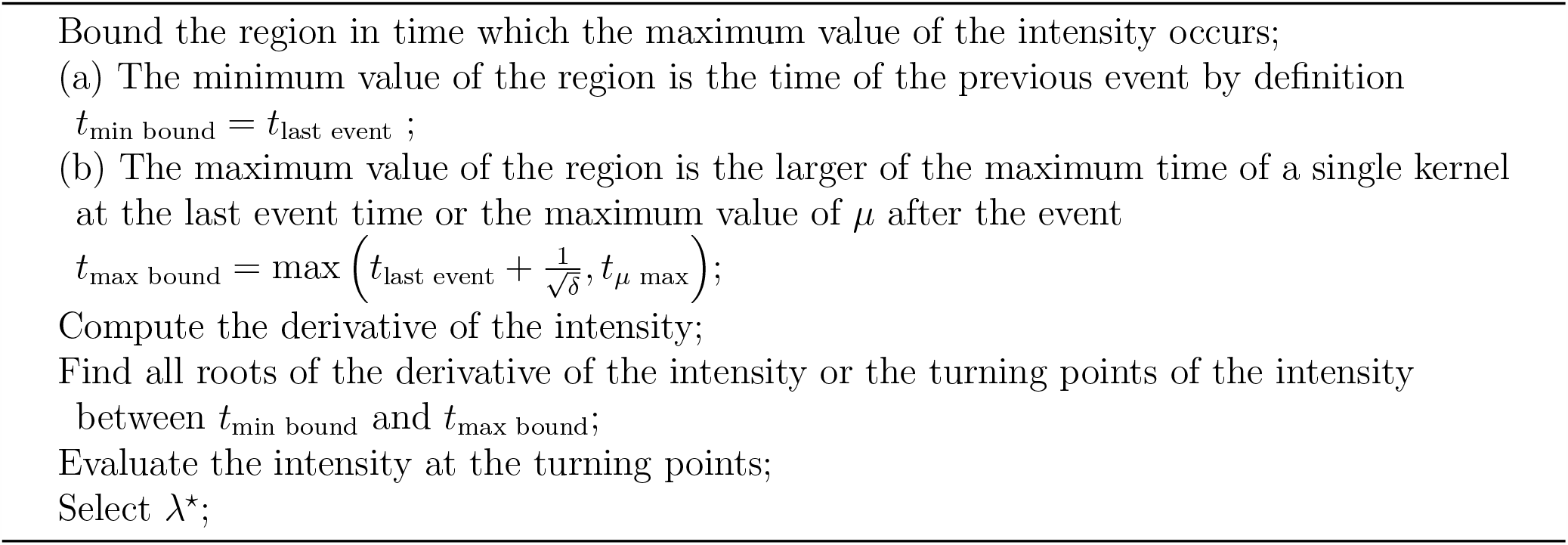

We simulated 10, 000 realisations of our Hawkes Process up to *T*_max_ = 1, 000 using Algorithms 1 and 2 and our fitted parameters. From this we could recreate the epicurve, or cumulative cases, over time. We also simulated 10, 000 realisations of just the *µ* term, or the endogenous cases only, which represented the imported malaria cases. We used the same algorithms as before, but set *α* = *δ* = 0 because we were not considering the cascade of infections from these importations at this time. We compared these simulations to a model fitted using the traditional exponential kernel with no delay.

It is also possible to use Hawkes Process models for prediction. We can see how well our model fits future data by not fitting our model to all the available data. Instead we hold back the last portion of the epicurve and forecasting over the period of the withheld data. We simulated for 35 days more than we fit to so that we could investigate the predictive power of the model. All our code is provided in the *epihawkes* package and available open source on GitHub^1^.

## 4 Results

We can recreate our kernel and exogenous term using the optimal parameters returned by our fitting procedure. Figure 2A shows the fitted serial interval for both China and Swaziland. The duration over which a person remains infectious, or where the intensity is greater than zero, is around 6 or 7 days for both China and Swaziland, but the individual contribution to the intensity from one person is greater in Swaziland than China. The kernel, *ϕ*(*t* − *t*_*i*_), is zero for the first 12 days, which corresponds to the delay in a person becoming infectious after they are bitten. A very different serial interval is obtained if the standard exponential kernel is used without a delay, see Appendix E. Figure 2B shows how *µ* varies over time for our proposed model. This variation is very different between China and Swaziland; *µ* decreases significantly over the 1000 days in China, but the initial intensity is much lower in Swaziland and increases slightly. Using these parameters, we calculate the *R*_*c*_ for China to be 0.299 and Swaziland to be 0.359.

**Figure 2:**
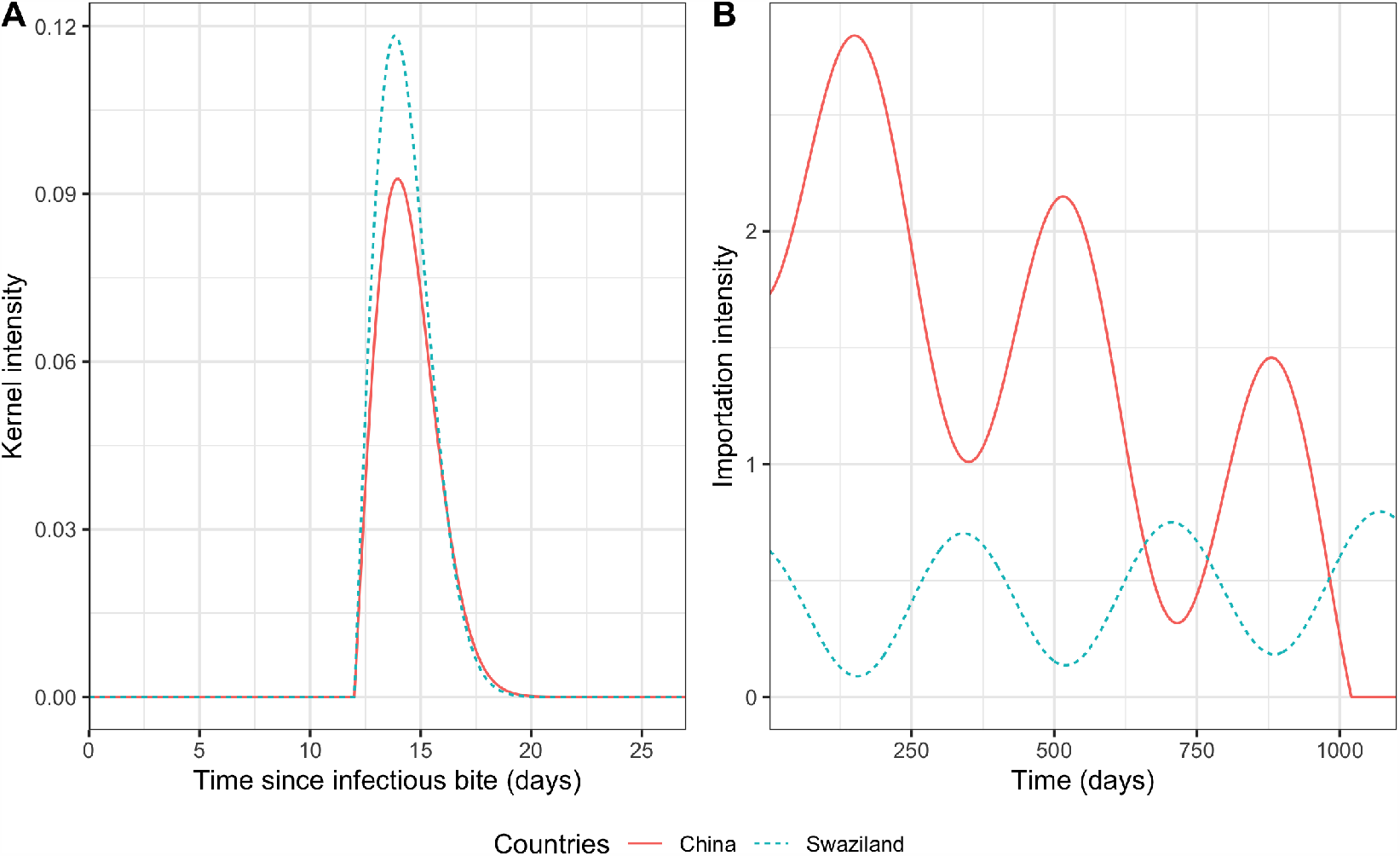
Fitted endogenous and exogenous terms for the China and Swaziland data. Figure A shows the fitted serial interval distribution for a single infection, which corresponds to the kernel given in equation (8). Figure B shows how the exogenous terms vary through time. The solid red lines correspond to the China data and the dashed blue lines correspond to the Swaziland data. Our fitted parameters correspond to

Our 10,000 simulations show different realisations of the Hawkes Process model and enable us to validate our fitting. Our intuition says that these simulations represent different ways that malaria could have transmitted in alternative scenarios. Figures 3A and C show malaria case counts over time for China and Swaziland respectively. The solid red line shows observed cumulative cases over time and the green line shows observed cumulative imported cases over time. Similarly, the black lines show cumulative cases from each simulation and the blue lines show cumulative imported cases from each simulation. There is very good agreement between the simulated data and the real case counts they are fitted to, especially in China where the red line lies within the bounds of our simulations. We are also able to separate out the cases which are importations from the ones that are from within country transmission, which is important in near-elimination settings. Figures 3B and D show the time varying intensity for China and Swaziland. The red line shows the intensity calculated using equation (1), the fitted parameters and the real event times. This agrees well with the simulated intensity calculated using the simulated event times and fitted parameters, which mostly bound the red line. We also simulated from a Hawkes Process model fitted with a exponential kernel, no delay and the exogenous term given in equation (9) - these simulations did not recreate the data well and had orders of magnitude more infections. Either too few infections were caused by importations and elimination occurred in the majority of scenarios or the intensity of the local infections was too long resulting in a sustained outbreak.

**Figure 3:**
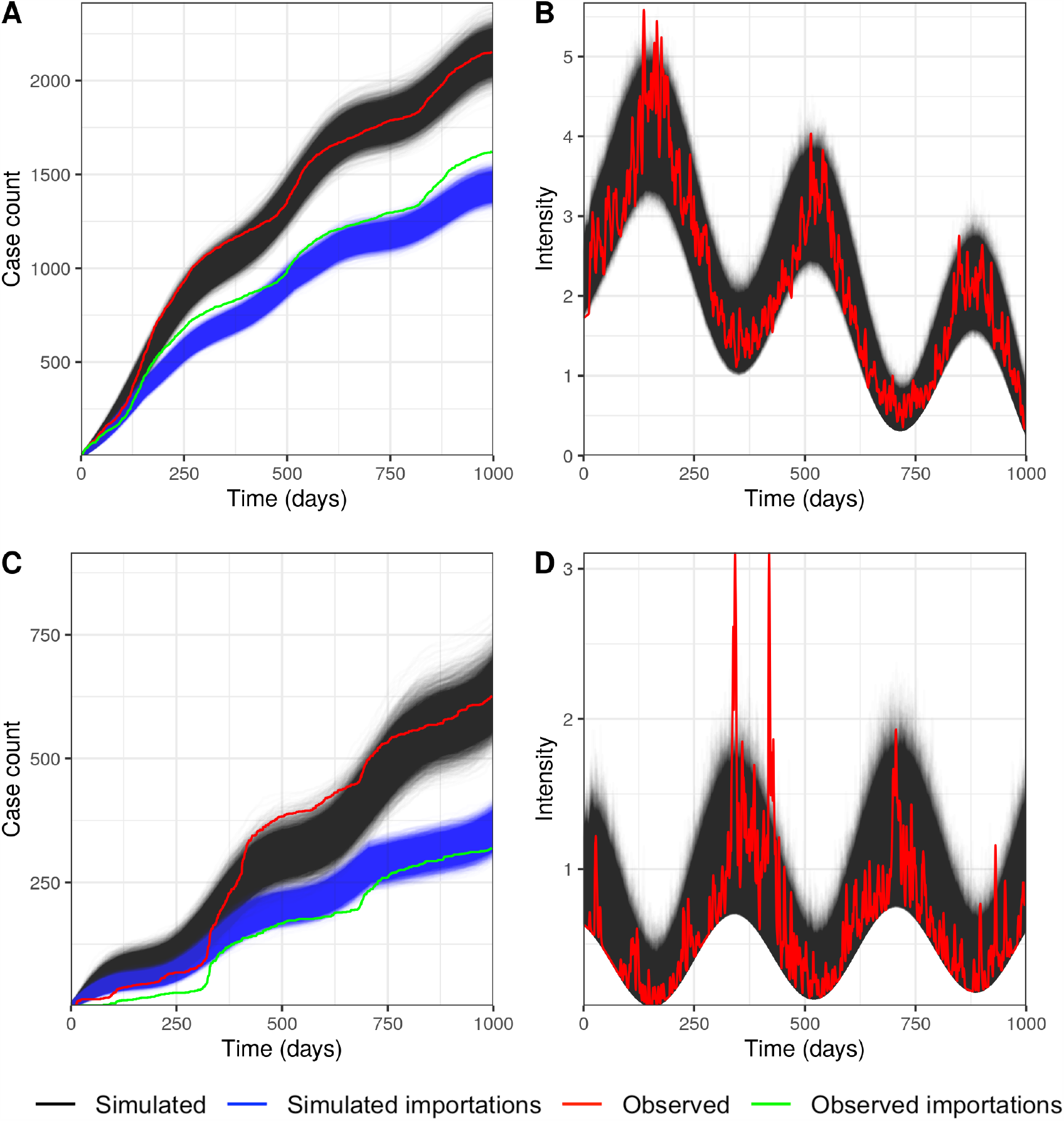
Simulated counts and intensities for the China and Swaziland data. Figures A and C show malaria case counts for China and Swaziland respectively. The red line shows the real case counts over time and the black lines show the case counts over time from 10,000 simulations of the full fitted model. The green line shows the real case count over time from the cases labelled as importations and the blue lines show the case counts over time from 10,000 simulations of just the exogenous term (equation (9)). Figures B and D shows the calculated Hawkes intensity (equation (1)) for China and Swaziland respectively. The red line shows the intensity calculated from the fitted parameters and real events, whereas the black lines show the intensity calculated from the fitted parameters and the simulated events.

It is also possible to use Hawkes Process models to predict future cases of malaria in a country. Figure 4 shows predicted cumulative cases of malaria over the next 35 days after we stop fitting our model. We get very good agreement between the real cases (red crosses) and the 10,000 simulations for approximately one month into the future. It is possible to predict further, but the predictions become less reliable. In particular in China, the fitted exogenous term has reached zero, meaning the simulations suggest that elimination has occurred. If we refit with more data, the *µ*(*t*) trend alters slightly and elimination is delayed.

**Figure 4:**
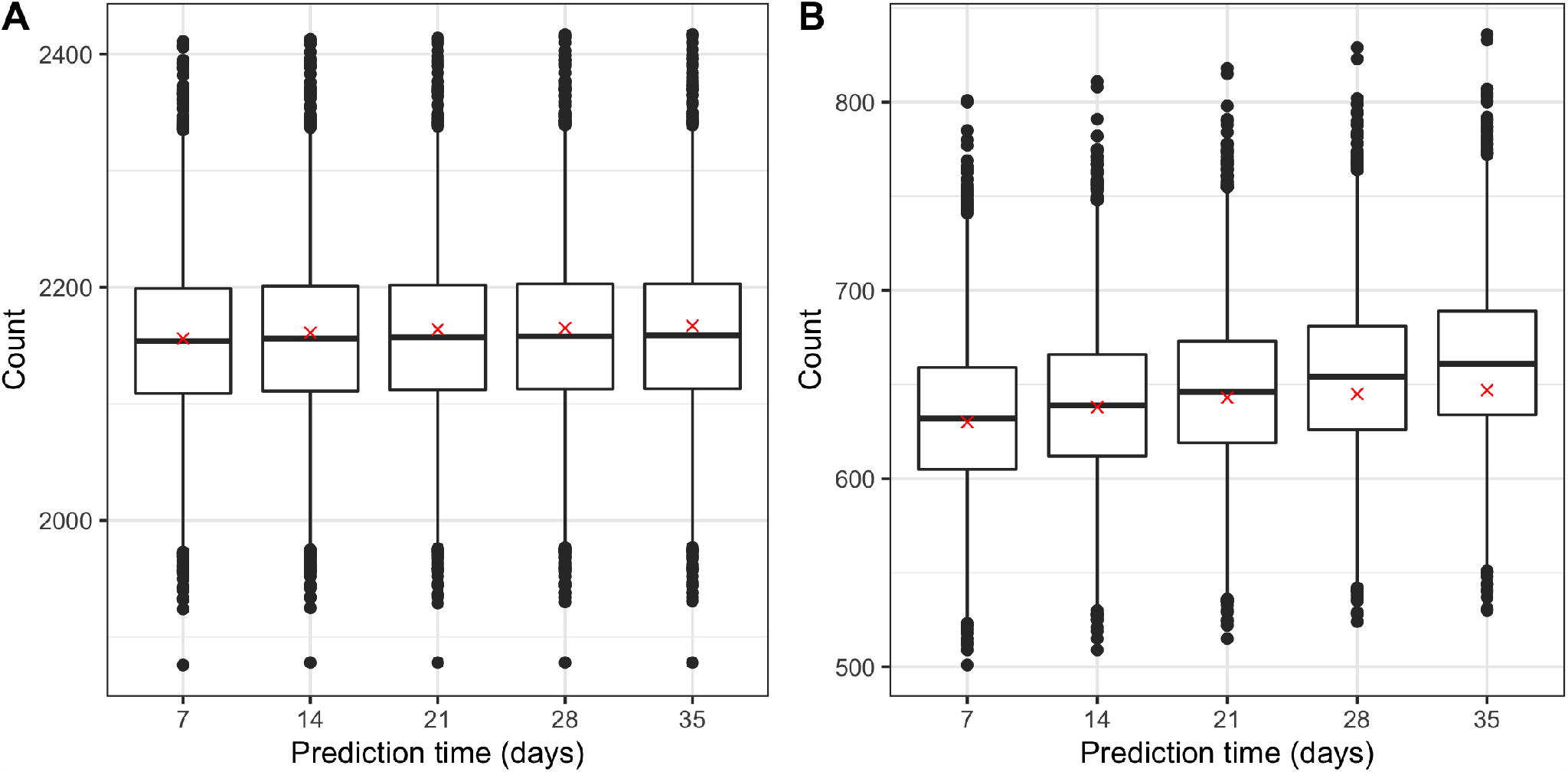
Predicted cumulative cases of malaria. Figure A shows weekly predicted cases of malaria for China and Figure B for Swaziland respectively. The red crosses show real cumulative cases and the box and whisker plot show predictions from the 10,000 simulations.

## 5 Discussion

Mathematical modelling is an important tool for helping countries close to eliminating malaria reach their goals. Recreating disease transmission patterns in low-endemicity settings is an important first step for validating these methods and their utility for informing policy. In this paper, we have shown that semi-mechanistic Hawkes Process models can be used to model the number of infections of malaria over time in both Yunnan Province, China, and Swaziland. We have also shown that it is necessary to make disease specific modifications to the traditional kernel to recreate malaria transmission. Our serial interval agrees with the 18 day intrinsic incubation period given in Reiner et al. [34] (approximately six days in the liver, which matches the duration in Figure 2A, plus the 12 day delay in the infectious period for mature gametocytes to be produced in sufficient densities). We also estimated similar case reproductive numbers as other methods using the same data. Routledge et al. [5] estimate a mean *R*_*c*_ of 0.29 in 2011, 0.25 in 2012 and 0.11 in 2013, which is similar to our estimate of 0.299 using a novel method. Similarly, Reiner et al. [34] estimate the *R*_*c*_ for Swaziland in different regions between 0.08 and 1.70, which encompasses our estimate of 0.359. These methods enable us to include mechanisms of transmission that are not considered in purely statistical methods but do not need the same quality of data that is necessary for network models. We do not capture the initial increase in cases between approximately days 350 and 400 well in Swaziland. Figure 2D suggests that the magnitude of our intensity function is not large enough to cause the number of secondary cases seen. However, Figure 2C shows there is also a spike in importations around this time, which our single exogenous term does not reproduce, and could be driving this increase in cases. This could reflect a policy that was changed during year 2 of the data, which decrease the number of importations in the subsequent years.

The use of Hawkes Processes is especially well suited to malaria modelling in near-elimination settings because not only can these methods be used to recreate cases over time, but they can be used to disentangle the relative contribution of importation verses local transmission. This is especially important in scenarios where *R*_*c*_ *<* 1 and malaria transmission transition from being community driven to being driven by importations. In these situations, understanding how many cases are being imported is perhaps more important to policy makers than the reproduction number, since local transmission is not sustained. Our fits to the overall case data are better than to our importations because we choose the parameters for the Hawkes Process that minimise the error in the cumulative case counts and do not include information about travel history or which cases were imported in our fitting procedure. We choose this parameterisation for our log-likelihood because we wanted to showcase how this method could be used to ascertain the proportion of imported malaria cases when the health systems do not know how many cases originated outside the community.

A benefit of modelling malaria transmission is that we can extend our models and forecast future behaviour. We show that in both China and Swaziland our median estimated case counts matches the actual case count very well. This is especially useful to enable policy makers to act appropriately to reduce the transmission. From Figure 2 we see that China has very successfully managed to reduce importations over the time period studied, whereas, importations have increased slightly during that study.

We recognise that despite this novel method providing a flexible and useful tool for modelling malaria there are several limitations. Our method requires a unique time stamp for each individual malaria case. This is often not available in the line lists provided by the surveillance system because they are recorded by the day of presentation of symptoms. We therefore add noise to the data to recreate unique timings. We investigated the impact of adding different types of uniform or normally distributed noise to our dates but this did not impact the fits of our model significantly. We also obtain very different optimum parameter sets depending where we start our optimisation algorithm from. This is because our parameter space is complex and there are many local maxima and minima. We compensate for this by starting our optimisation algorithm from many different places to ensure, that although we may not find the “true” optimum we achieve a good fit to the data and our values for *R*_*c*_ are in line with other findings in literature. We only consider a snapshot of dates because we want to compare our forecasts of the model to true data and simulation is slow because we are solving a NP hard problem to find the maximum intensity of the Rayleigh kernel with a delay. Speeding this up is an area of ongoing research.

## Data Availability

Code is available at https://github.com/mrc-ide/epihawkes.

## Acknowledgements

The authors would like to thank Joshua Proctor for early discussions about using Rayleigh kernels to model malaria and for his comments on the final draft. HJTU is funded by Imperial College London through an Imperial College Research Fellowship grant. HJTU, SM, IR and SB acknowledge joint centre funding by the UK Medical Research Council (MRC) and the UK Department for International Development (DFID) under the MRC/DFID Concordat agreement and is also part of the EDCTP2 programme supported by the European Union.

## A Hawkes Likelihood

Following Rasmussen [39] and Rizoiu et al. [23], the likelihood of the Hawkes process can be derived as follows. We define the history, *H*_*t*_, of the disease outbreak to be the list of infection times *t*_1_, *t*_2_,, *t*_*n*_up to but not including the current time *t*. We also define the conditional intensity function (or hazard function) in terms of time *t* as

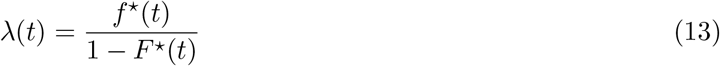

where *f*^⋆^(*t*) := *f* (*t*|ℋ_*t*_) is the conditional probability density function of the time of the next event *t*_*n*+1_ given the history of the previous events *t*_1_, *t*_2_, …, *t*_*n*_ and *F*^⋆^ (*t*) is the corresponding cumulative distribution function. Equations for *f*^⋆^(*t*) and *F*^⋆^(*t*) are derived as follows.

We first write the conditional intensity function, equation (13), just in terms of the cumulative distribution function *F*^⋆^(*t*)

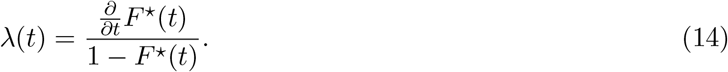

We then simplify equation (14) into

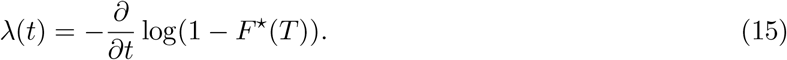

If we integrate equation (15) between times *t*_*n*_ and *t* we get

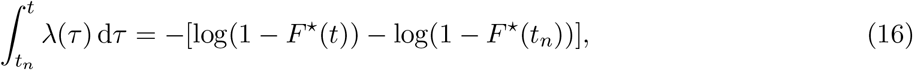

where *t* is the current time and *t*_*n*_ is the time of the last known event prior to *t*. Since *F*^⋆^(*t*_*n*_) = 0, because *t*_*n*+1_ > *t*_*n*_, equation (16) can be simplified to be

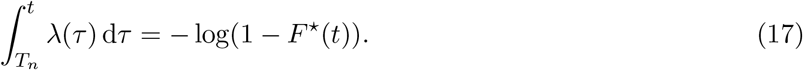

Rearranging equation (16) gives us the equation for the cumulative distribution function

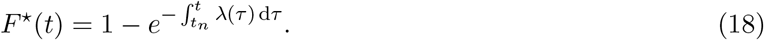

Substituting equation (18) into equation (13) enables us to solve for the conditional probability density function

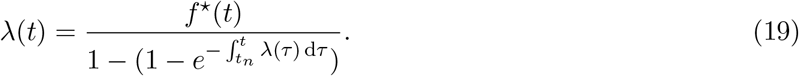

After rearranging equation (19),

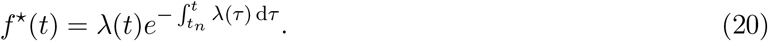

The likelihood function of the Hawkes Process, with parameters *θ* is the joint density function of all the points in the history of the outbreak and can therefore be factorised into all the conditional densities of each points given all points before it. This yields

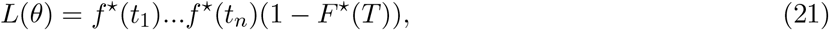

where (1 − *F*^⋆^(*T*)) is the last term because the unobserved point *t*_*n*+1_ appears after the end of the observation interval. Using equation (13), the likelihood function can be written as

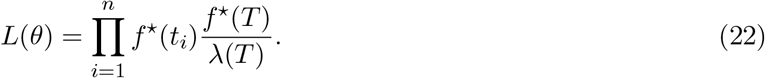

Expanding equation (22) using the equation for the conditional probability density function from equation (20) gives

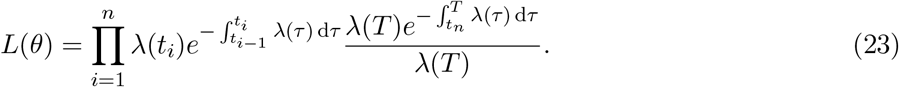

Simplifying equation (23) yields

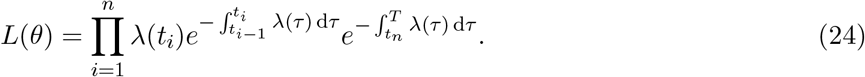

By combining the exponentials in equation (24) and assuming *t*_0_ = 0, the likelihood function is

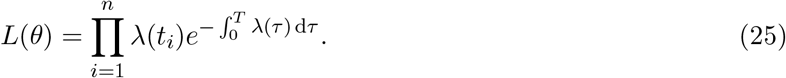

One method for selecting parameters is to maximising our likelihood function over our parameter space Θ, which is defined as

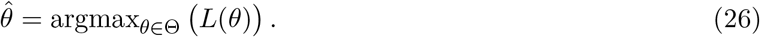

However, It is common to minimise the negative log-likelihood function instead of maximising it because it is a less computationally expensive calculation and is more accurate. We therefore define our problem as

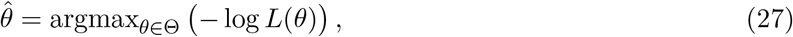

Where

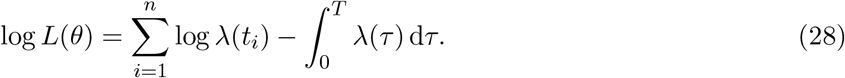

## B Directional derivatives of the negative log-likelihood

Our fitting process is defined as

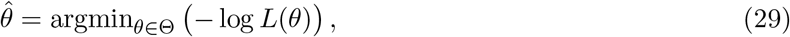

for

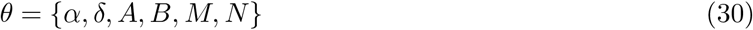

where

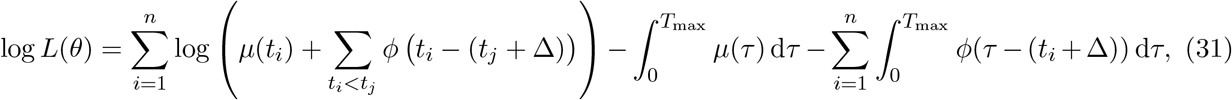

and *ϕ*(*·*) is the Rayleigh kernel in equation (8) and *µ*(*t*) is given in equation (9). We define the integral of *µ* with respects to *t* between 0 and *T*_max_ to be

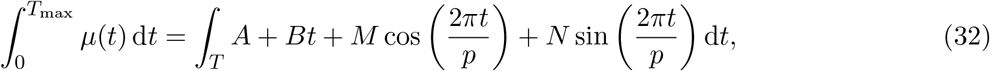

where

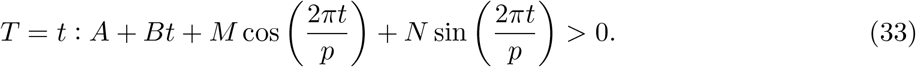

Assuming that there are a finite number of regions between 0 and *T*_max_ where 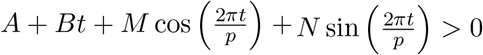 and each region is between [*a*_*k*_, *b*_*kj*_],

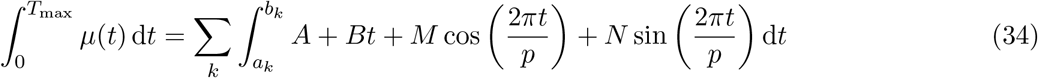

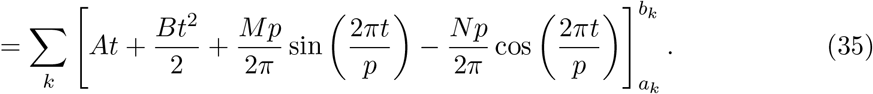

Therefore, in full

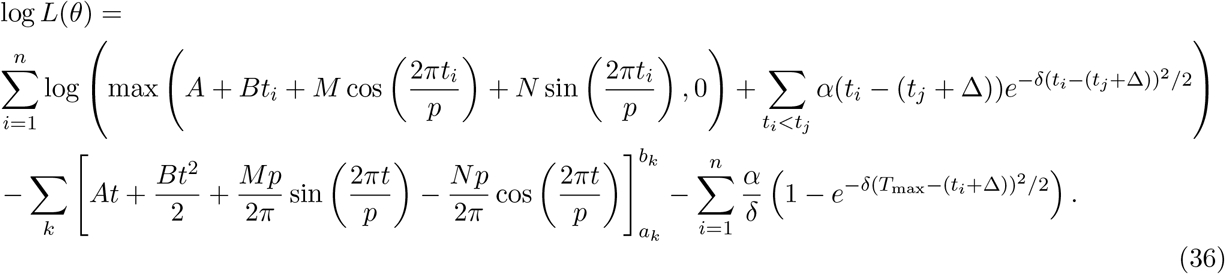

We need directional derivatives of the likelihood with respect to each parameter as an input for the optimisation algorithm. These are written out below for completeness. We define

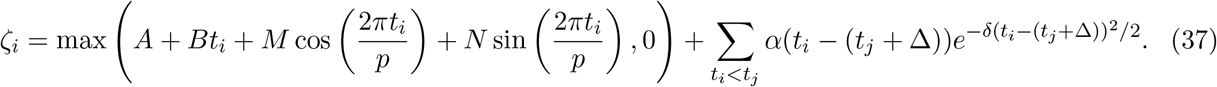

First, we consider the directional derivatives for the kernel parameters:

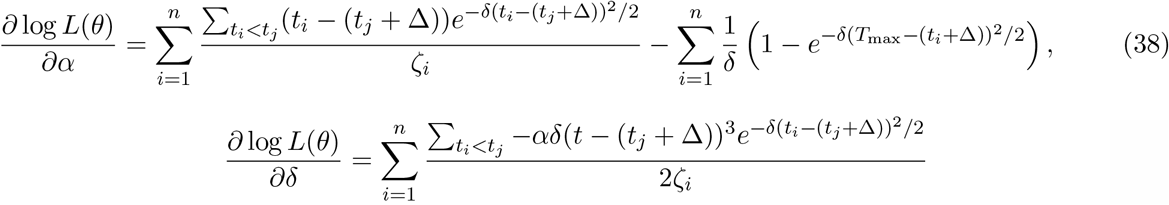

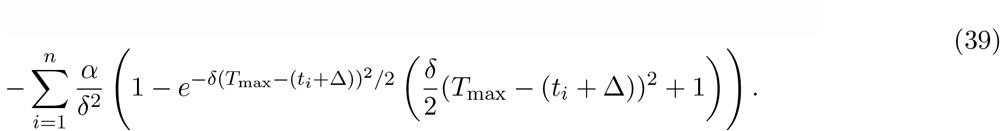

Then we consider the directional derivatives for the *µ* parameters; the first term of each directional derivatives is only evaluated at times, *t*_*i*_, when *µ*(*t*_*i*_) > 0:

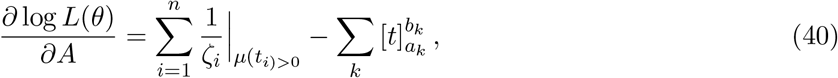

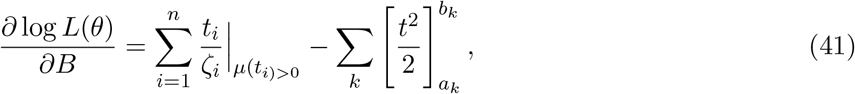

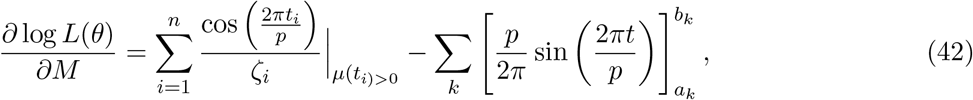

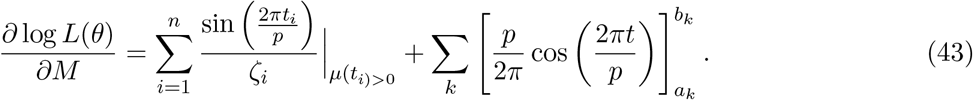

The derivative of *µ* with respects to *t* is required for the simulation so included here for completeness

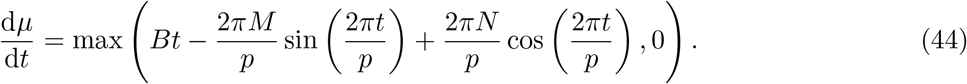

## C Maximum of intensity using Rayleigh kernel

There is no analytic solution for the time of the maximum value of the intensity function, *λ*_max_, after the last event or infection so it is no longer trivial to find the maximum value of the intensity. We are however able to bound the region of time in which this maximum lies by considering the superposition of multiple Rayleigh kernels and the functional form of the exogenous term *µ*. We consider *µ* = 0 in this appendix and present the results when *µ* varies in Section 3.2. First we assume there is no delay in a person becoming infectious once they have been infected and then consider a delay.

### C.1 Rayleigh kernel without a delay

The value of the intensity function when *µ* = 0, or is constant, can be found by superposing the Rayleigh kernels that result from individuals being infected at different times. Figure C.5 shows examples of the intensity functions for events at different times with kernel parameters *α* = 1.0 and *δ* = 1.0. The time of the maximum value of the intensity for a single infection, *t*_max intensity_, is known see Figure C.5A. However, as shown in Figure C.5B and C, *t*_max intensity_ from two infectious does not occur at the same time as the maximum of either individual infection, so we can only place a bound around the time of *λ*_max_. The maximum value of the intensity after an event must occur after the last event by definition, so this forms the lower bound on the time region in which the maximum lies. Since we assume the parameters of the kernels are constant with time, all the kernels must decrease after the maximum value of the last event so the upper bound must be the maximum time of a Rayleigh kernel that starts at the time of the last event. Therefore,

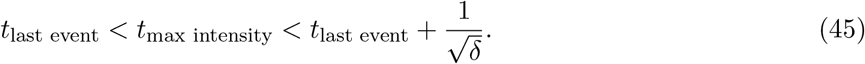

This is shown in Figures B and C for two events because the solid black line is between the blue dot and dashed blue line and again shown in Figure D for 6 events since the black line lies between the dark red dot and dark red dashed line.

Now we have the bound in which *t*_max intensity_ lies, it is possible to use a root finding algorithm to locate *t*_max intensity_ and thus *λ*_max_. We know *λ*_max_ is a turning point in *λ*(*t*), so *t*_max intensity_ is the root of the derivative of *λ*_max_. This is analytically

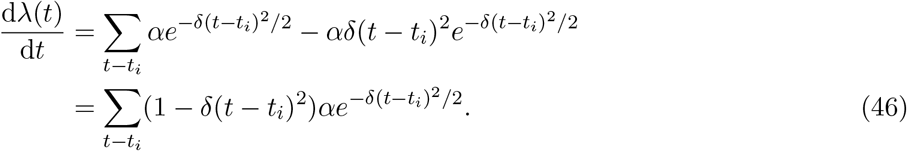

**Figure C.5:**
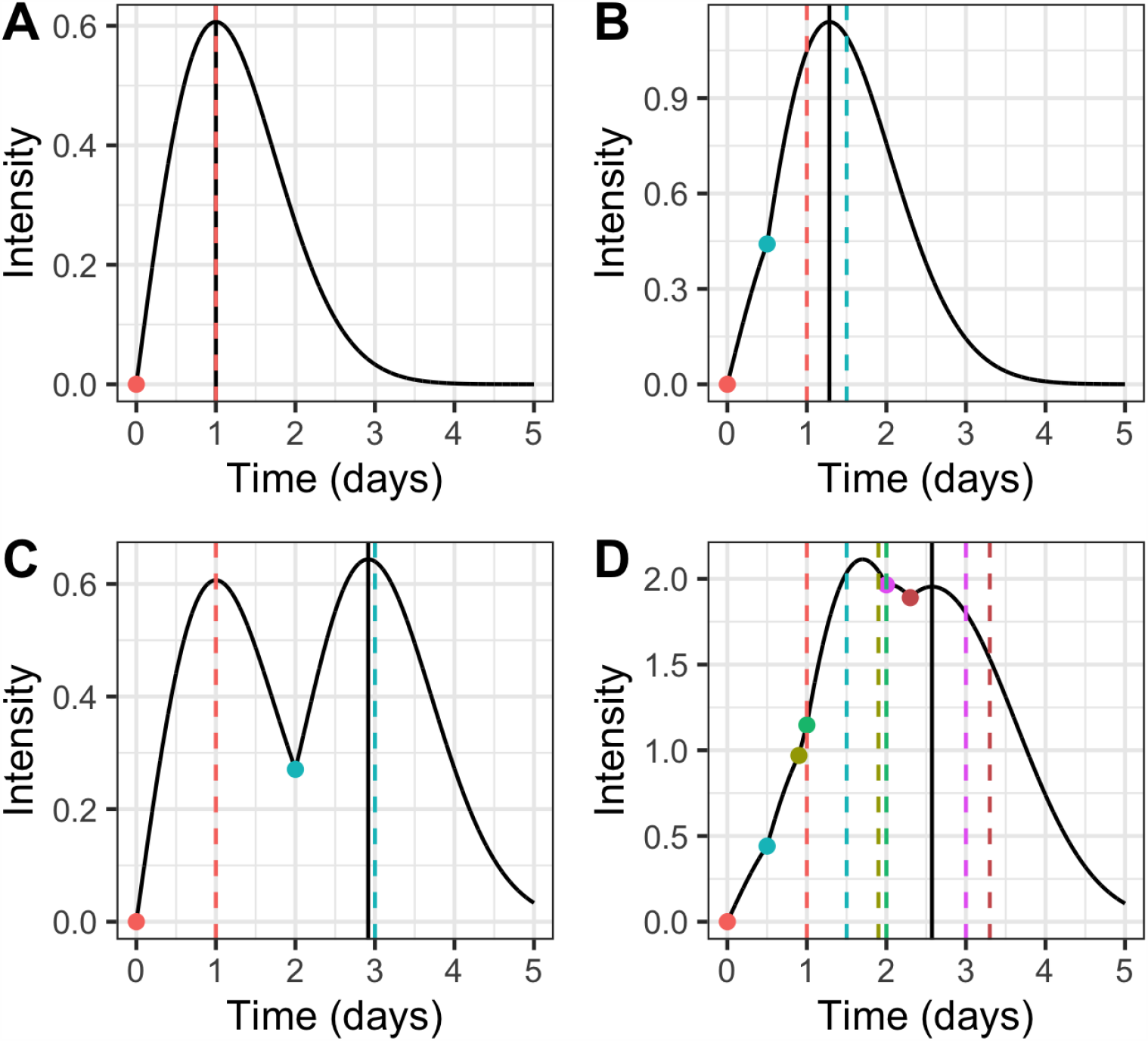
Intensity function for events at different times with kernel parameters *α* = 1.0 and *δ* = 1.0. The coloured dots refer to different infections and the corresponding coloured dashed lines indicate the time of the theoretical maximum value of a single Rayleigh kernel at each event time. The solid black line indicates the time of the maximum value of the kernel after the last event, which results from the summing the contribution from the Rayleigh kernel from each event. Figure A shows a single Rayleigh kernel from one infection, Figures B and C show the superposition of two infections and Figure D shows the superposition of 6 infections.

**Figure C.6:**
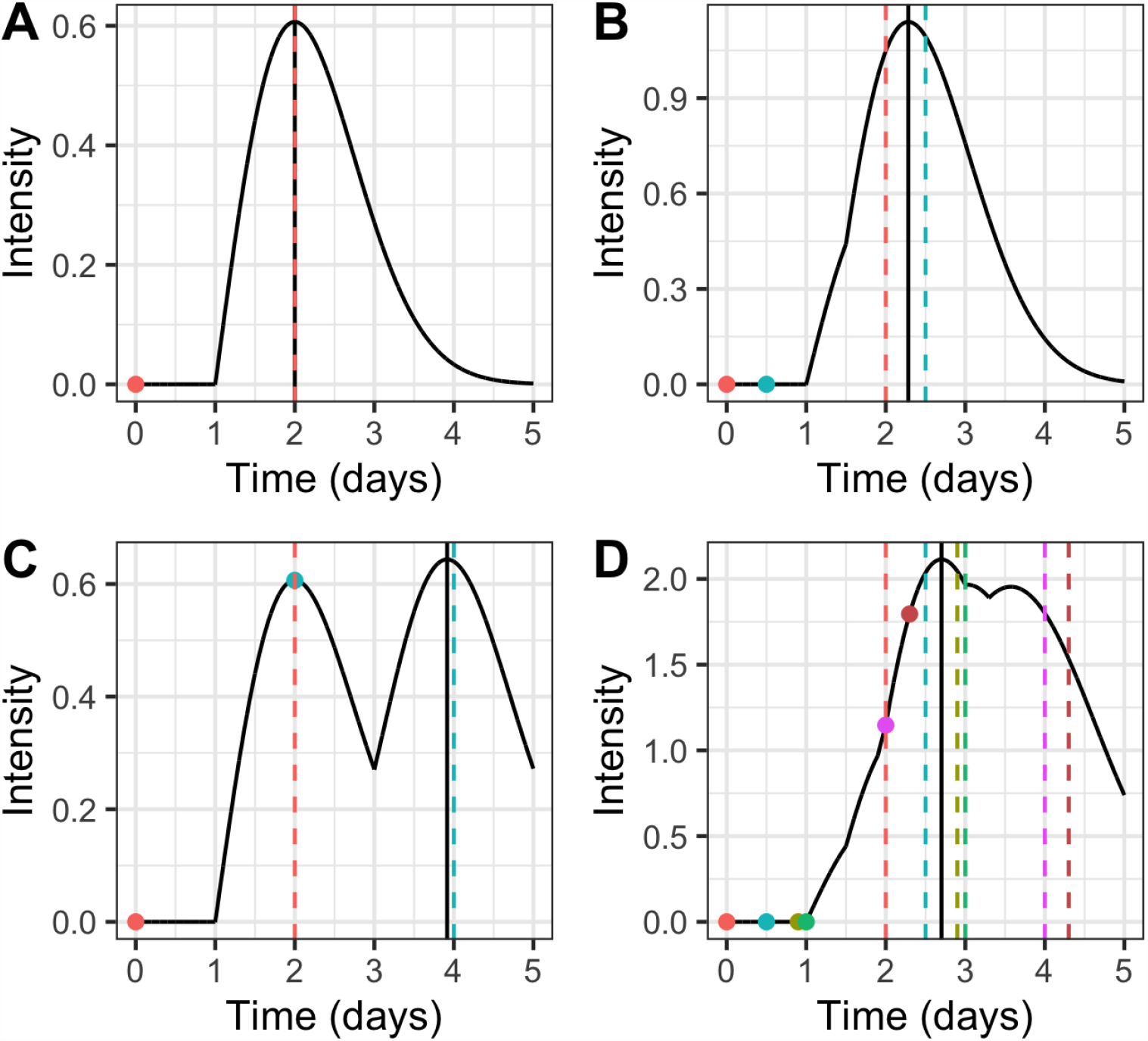
Intensity function for events at different times with kernel parameters *α* = 1.0 and *δ* = 1.0 and a 1 day delay. The coloured dots refer to different events or infections and the corresponding coloured dashed lines indicate the time of the theoretical maximum value of a single Rayleigh kernel at each event time. The solid black line indicates the time of the maximum value of the kernel after the last event. Figure A shows a single Rayleigh kernel, Figures B and C show the superposition of two events and Figure D shows the superposition of 6 events.

## C.2 Rayleigh kernel with a delay

When we add a delay, there is no longer necessarily only one root between *t*_last event_ and 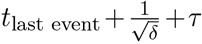, where *τ* is the delay. Therefore, using root finding algorithms for the derivative may only locate a local maximum and not the true global maximum of the intensity, which is needed for the simulation algorithm. Figure C.6 shows examples of the intensity function for the same scenarios as Figure C.5, but includes a 1 day delay before a person becomes infectious. Again, in all Figures the maximum value of the kernel lies between the time of the last event and the time of the maximum value of a single kernel at that time but in Figure D it is obvious that there are three local maximum in that range so care needs to be taken to select the true maximum. This can be done by evaluating all the roots of the derivative of *λ*(*t*) between *t*_last event_ and 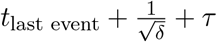 and then selecting the maximum value.

## D Branching factor

The branching factor, *n*^⋆^, is defined as the expected number of events that are directly triggered by an event in a Hawkes process. It is defined as

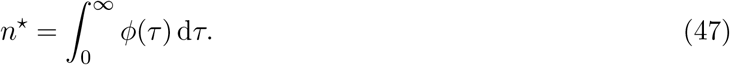

Substituting equation (7) into equation (47), and assuming *τ* = *t* − *t*_*i*_ results in

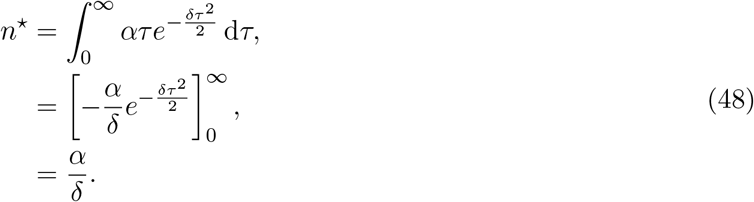

**Figure E.7:**
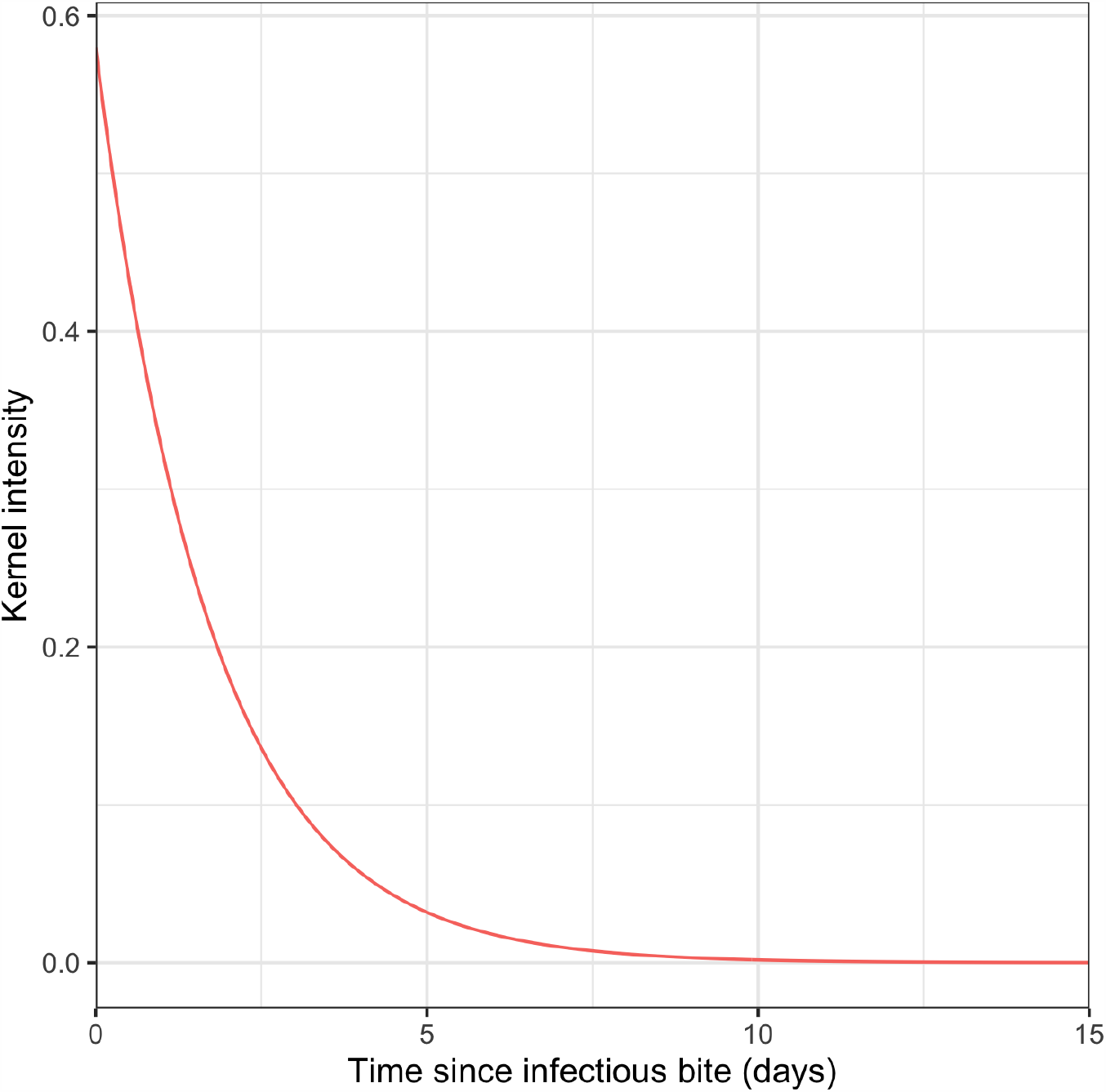
Fitted endogenous for China using a standard exponential kernel without a delay,

## E Fitted exponential kernel

## Footnotes

1 https://github.com/mrc-ide/epihawkes

